# Contrasting impacts of social deprivation and ethnicity on novel vs. established cardiovascular procedures: A population-level study of TAVR and SAVR

**DOI:** 10.1101/2025.03.17.25324150

**Authors:** Hasrit Sidhu, Feng Qiu, Ragavie Manoragavan, Derrick Y. Tam, Maneesh Sud, Andrew Czarnecki, Mamas A Mamas, Harindra C. Wijeysundera

## Abstract

**Background:** Social deprivation markers are associated with worse clinical outcomes after cardiac revascularization procedures. However, the association between social deprivation and cardiac valvular procedural outcomes is less established. We sought to distinguish the relationship between social deprivation and outcomes in the management of severe aortic stenosis, specifically transcatheter aortic valve replacement (TAVR) versus conventional surgical aortic valve replacement (SAVR).

**Methods:** Demographics, patient comorbidities, procedural details, and outcomes for adults undergoing TAVR and SAVR between April 2017 and March 2022 were obtained from clinical and administrative databases and linked to neighbourhood-level measures of social deprivation using the Ontario Marginalization Index (ON-MARG) in Ontario, Canada. The three dimensions of social deprivation assessed were (1) material deprivation, (2) residential instability and (3) ethnic concentration. Our outcomes were 30-day mortality, 30-day readmission, 1-year mortality and 1-year readmission. Separate Cox proportional hazard models for post-procedural mortality and cause-specific hazard models for post-procedural re-admission were used to determine the association between social deprivation and post-procedural outcomes after TAVR versus SAVR.

**Results:** We identified a total of 6,218 TAVR procedures and 3,342 isolated SAVR procedures within our study period after exclusion criteria were applied. After multivariable adjustment, we found that TAVR was associated with lower 30-day mortality (HR 0.58; 95% CI [0.37, 0.92]; p = 0.02), lower 30-day readmission rates (HR 0.75; 95% CI [0.63, 0.89]; p-value = 0.001) and a higher 1-year readmission rate (HR 1.14; 95% CI [1.02, 1.27]; p-value = 0.01) when compared to SAVR. When the three ON-MARG domains by treatment interactions were included in the analysis, the associations between TAVR and SAVR and these outcomes were not modified by the degree of neighbourhood social deprivation.

**Conclusion:** TAVR is associated with lower 30-day mortality and 30-day re-admission rates and higher 1-year re-admission compared to SAVR. These associations were not modified by social deprivation, including ethnic concentration, material deprivation and residential instability.

## INTRODUCTION

Over the past decade, landmark trials have shown both non-inferiority and superiority of TAVR over SAVR for the treatment of severe aortic stenosis in high-risk, intermediate-risk and low-risk surgical patients^1–4^. This has led to the clinical indications for TAVR expanding significantly over this timeframe. However, timely referral, access to care, and post-procedural outcomes may be impacted by social determinants of health.

Social deprivation in health care is defined as the reduction of normal interaction between an individual and their associated healthcare system^5^. It is characterized by one’s ethnicity, residential instability and economic status. Literature over the past decade has delineated the impact of social deprivation on outcomes after cardiac procedures, such as percutaneous coronary intervention (PCI), TAVR, coronary artery bypass grafting (CABG) and SAVR. The relationship between social deprivation and an increased length of hospitalization and mortality after PCI and cardiac surgery is well established^6,7^. Residential instability is known to be associated with an increased risk of mortality and re-admission 1-year post-TAVR^8^. Additionally, ethnic marginalization in the United States is associated with increased mortality rates after SAVR, however, the relationship between ethnicity and outcomes after TAVR remains less clear^9,10^. Moreover, there is no data comparing the outcomes of these procedures and their association with other markers of social deprivation, including material deprivation and residential instability, which both may represent a complex interplay with ethnicity. Most importantly, it is unknown if the relationship between social deprivation and post-procedural outcomes is different in procedures such as TAVR that are less established compared to traditional surgical alternatives, such as SAVR.

Accordingly, to address this knowledge gap, we sought to utilize the ON-MARG, a multi-dimensional metric of neighbourhood-level social deprivation to study the relationship between social deprivation and mortality and re-hospitalization after SAVR and TAVR, using a population-based study in Ontario, Canada. This knowledge will provide an initial insight into the interplay between social deprivation and clinical outcomes after novel and established cardiac procedures and inform future work with regards to the adoption of emerging therapies.

## METHODS

### Transparency and Openness Promotion Statement

The data that support the findings for this study are not available given restrictions based on privacy regulations in Ontario, Canada.

### Study design and setting

This population-level study was conducted using administrative and clinical datasets located and analyzed at ICES (previously the Institute for Clinical and Evaluative Sciences) and linked using unique encoded patient identifiers. ICES is an independent, not-for-profit research institute with a wide range of Ontario health data, including anonymous patient records and multiple clinical databases. ICES is a prescribed entity under Ontario’s Personal Health Information Protection Act, allowing researchers to link de-identified population-based data with clinical registries to conduct approved research studies under regulated privacy and security policies and procedures (see link to Data and Privacy at www.ices.on.ca). The use of ICES data in this project was authorized under section 45 of Ontario’s Personal Health Information Protection Act, which does not require review by a Research Ethics Board and the need for individual patient consent was waived. Therefore, institutional review board approval was waived. We adhered to the Strengthening the Reporting of Observational Studies in Epidemiology (STROBE) statement for reporting of observational studies.

### Context

This study was conducted in Ontario, Canada, which is Canada’s largest province with a population of approximately 15 million people. All Ontario residents receive universal health care from a single third-party payer, the Ministry of Health. TAVR has been funded since 2012 and is available for all patient risk categories at the 11 cardiac centres in the province.

### Data sources

The primary data source was the CorHealth Ontario Registry, which contains information on all invasive cardiac procedures performed in the province. Reporting to the CorHealth Ontario registry for cardiac procedures, including TAVR and SAVR is a mandatory prerequisite for funding. This registry contains data on patient demographics, co-morbidities and procedural details. Its accuracy has been previously validated using retrospective chart review and comparisons with other databases^11^.

The CorHealth Ontario registry was linked to administrative databases at ICES including the Canadian Institute of Health Information (CIHI) Discharge Abstract Database (DAD) which was used to ascertain subsequent hospital readmission and supplement baseline patient comorbidities. Validated ICES-derived databases were used to identify diabetes, heart failure, hypertension, chronic obstructive pulmonary disease (COPD), and dementia. The Registered Persons Database (RPDB) was used for additional demographic variables including quintile of median neighbourhood income and rural residence, as well as vital status. The hospital frailty score was calculated for each patient using administrative datasets.

### Study population

We included all patients > 18 years who underwent TAVR or SAVR between April 1st, 2017, and March 31st, 2022 through the CorHealth Ontario Registry. All patients with prior cardiac surgery were excluded. For SAVR, only patients with a primary diagnosis of aortic stenosis were included (ICD codes I350, I352). If patients received both procedures, only their first procedure was included in the analysis. Concomitant procedures, such as concurrent CABG, mitral valve repair/replacement, tricuspid valve repair and aortic root repair/replacement with SAVR, were excluded from the analysis. Patients undergoing SAVR with a prior history of TAVR were also excluded from the analysis. The maximum follow-up date was March 31st, 2023. Patients with an invalid ICES key number (IKN) were excluded.

### Outcomes

Our main outcomes of interest were all-cause 30-day mortality, 30-day hospitalizations, 1-year mortality and 1-year hospitalizations after SAVR and TAVR. For mortality, outcomes were calculated from the day of the procedure. Readmission was counted after discharge from index hospitalization in the first 30 days or 1 year. Subsequent readmissions were not included in the analysis. Patients who died before discharge were censored from the re-hospitalization analysis. Data regarding death and hospitalizations were obtained from the CIHI-DAD and RPDB.

### Social deprivation

Neighbourhood-level data (using the 20,160 dissemination areas in Ontario as the geographic unit of analysis) on social deprivation was gathered from the 2016 version of the ON-MARG^5^. A dissemination area is a small, and stable geographic unit composed of one or more dissemination blocks with an average population of 400 to 700 persons based on data from the prior Census of Population Program. The ON-MARG is a geographical-based index derived from census data that can be used to measure health inequities in Ontario across four dimensions: *dependency* (concentrations of individuals having no income including seniors, children, and adults whose work is not compensated), *material deprivation* (income, quality of housing, educational attainment and family structure characteristics), *ethnic concentration* (recent immigrants, and/or belonging to a visible minority group as defined by Statistics Canada), and *residential instability* (types and density of residential accommodations, and family structure characteristics) (Table 1). For the purpose of our study, we only evaluated three of the dimensions, namely material deprivation, ethnic concentration and residential instability. Dependency is strongly co-linear with age, which is a known driver of procedural outcomes; thus, we excluded dependency from our analysis.

**Table 1:**
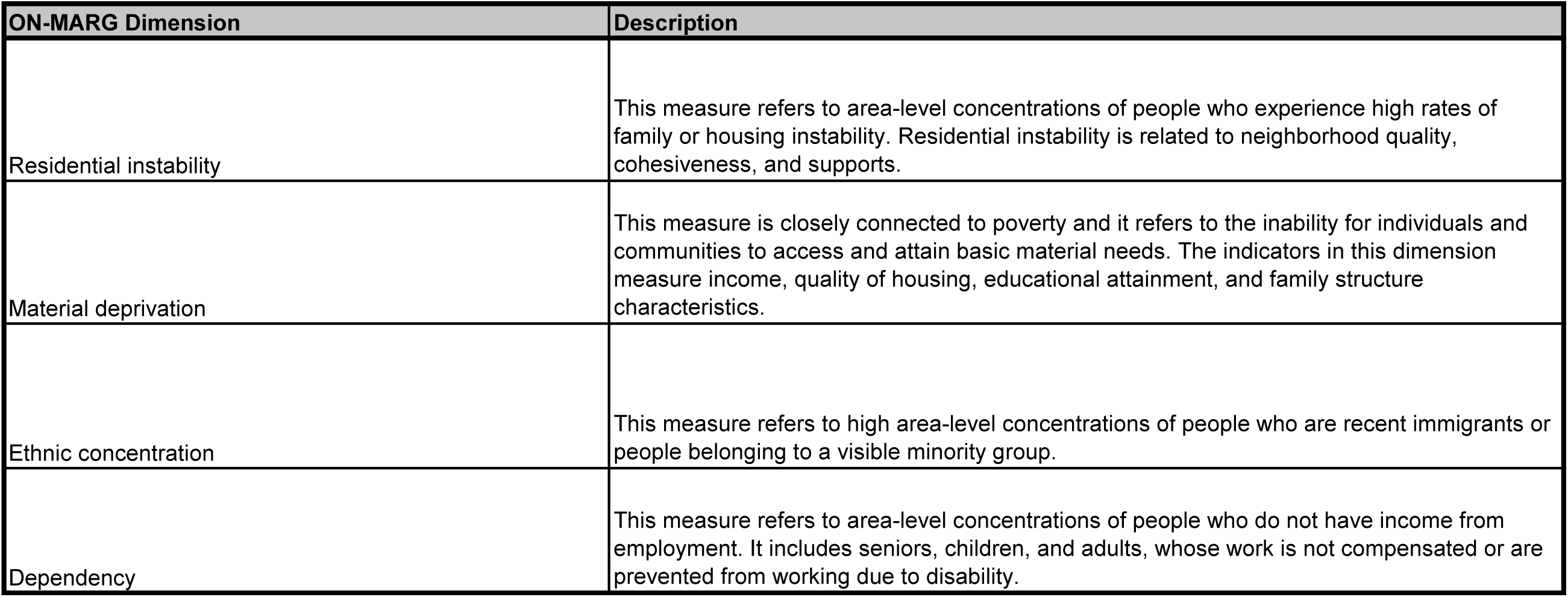
Description of ON-MARG Markers of Social Deprivation. *ON-MARG indicates Ontario Marginalization Index*.

| ON-MARG Dimension | Description |
| --- | --- |
| Residential instability | This measure refers to area-level concentrations of people who experience high rates of family or housing instability. Residential instability is related to neighborhood quality, cohesiveness, and supports. |
| Material deprivation | This measure is closely connected to poverty and it refers to the inability for individuals and communities to access and attain basic material needs. The indicators in this dimension measure income, quality of housing, educational attainment, and family structure characteristics. |
| Ethnic concentration | This measure refers to high area-level concentrations of people who are recent immigrants or people belonging to a visible minority group. |
| Dependency | This measure refers to area-level concentrations of people who do not have income from employment. It includes seniors, children, and adults, whose work is not compensated or are prevented from working due to disability. |

The degree of marginalization was quantified by quintiles. Quintiles describe marginalization for each dimension on a scale of one to five for each geographic unit, with one being the least marginalized and five being the most marginalized.

### Statistical Analysis

We developed cox proportional hazard models for post-procedural mortality and cause-specific hazard models for post-procedural readmission to account for the competing risk of death. Separate models were created for each ON-MARG dimension of ethnic concentration, residential instability and material deprivation. All models were adjusted for demographic risk factors (age, sex, and rural status), medical co-morbidities (frailty, heart failure, coronary artery disease, arrhythmias, peripheral vascular disease, cerebrovascular disease, diabetes, hypertension, dyslipidemia, chronic obstructive pulmonary disease, renal disease, dialysis, major neurocognitive disorder, liver disease, cancer and interstitial lung disease), prior coronary percutaneous intervention and fiscal year of the procedure. Adjusted outcomes were first compared between TAVR and SAVR. To determine whether social deprivation indices impacted the treatment assignment differently, an interaction term was added for treatment (i.e. TAVR/SAVR) and social deprivation. A robust variance estimator was used to account for clustering at the level of the dissemination area. A p-value of <0.05 was considered statistically significant.

## RESULTS

### Cohort

After applying appropriate exclusion criteria, we identified 6,218 TAVR procedures and 3,342 SAVR procedures in Ontario from April 1st, 2017, to March 31st, 2022 (Figure 1). Baseline characteristics are presented in Table 2. The mean age was 76.9 years for all patients. The TAVR cohort had a mean age of 81.7 years, while the SAVR cohort had a mean age of 68.0 years; 43.9% of all patients were female, with TAVR representing a greater distribution of female patients at 46.5% compared to 39% for SAVR. The clinical frailty score and Charlson score for medical comorbidities were higher, as expected, for TAVR compared to SAVR.

**Figure 1:**
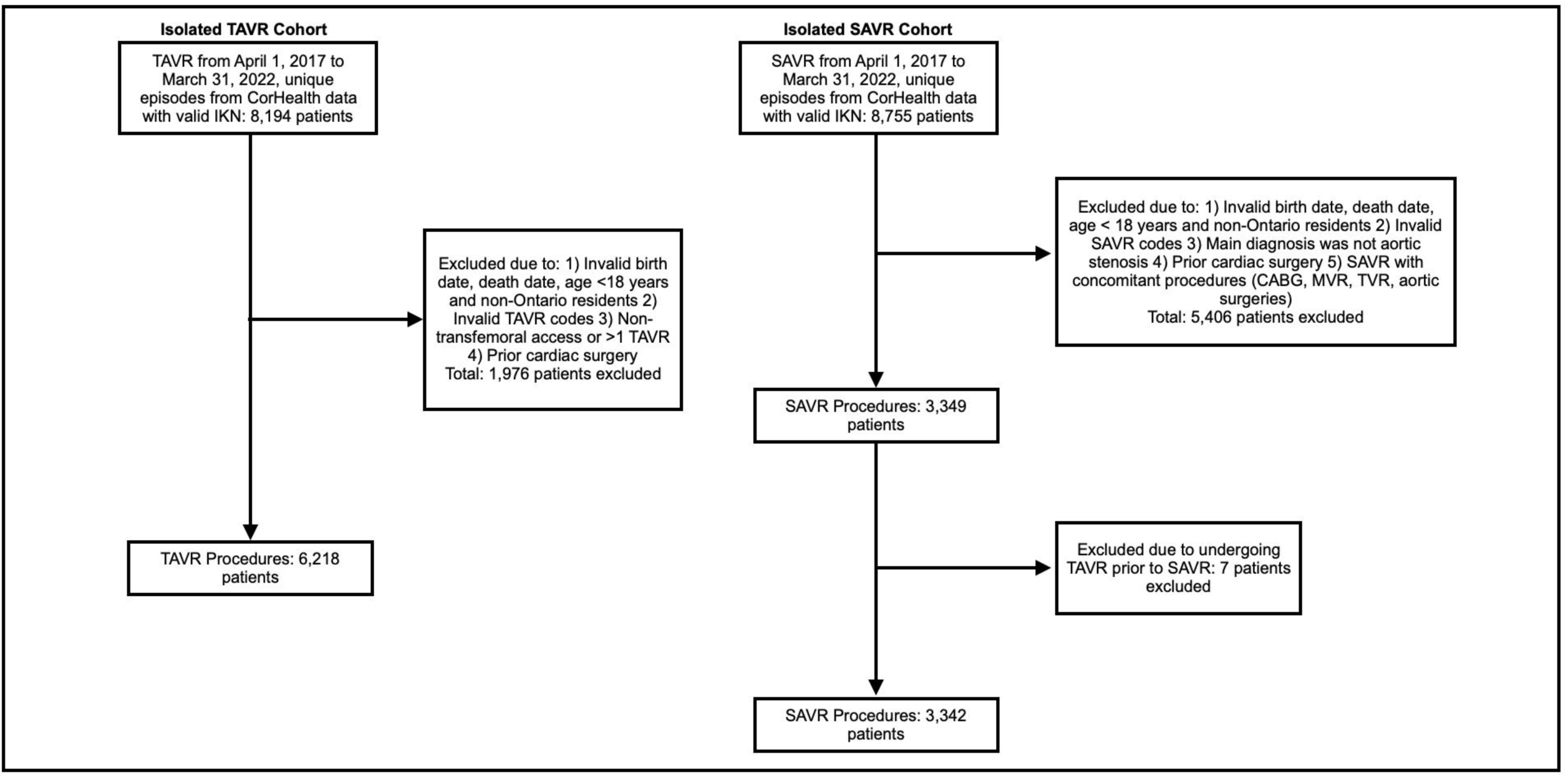
Distribution of all transcatheter aortic valve replacements (TAVR) and surgical aortic valve replacement (SAVR) procedures. */KN indicates ICES key number*.

**Table 2:** Baseline patient characteristics and medical comorbidities. TAVR, transcatheter aortic valve replacement SAVR, surgical aortic valve replacement SD, standard deviation

| Patient Characteristics | SAVR (N=3,342) | TAVR (N=6,218) | Total (SAVR & TAVR)<br>(N=9,560) |
| --- | --- | --- | --- |
| Age at referral, years<br>(mean ± SD) | 68.0 ± 9.3 | 81.7 ± 7.3 | 76.9 ± 10.4 |
| Female Sex | 1,304 (39.0%) | 2,893 (46.5%) | 4,197 (43.9%) |
| Rural | 607 (18.2%) | 717 (11.5%) | 1,324 (13.8%) |
| **Medical Comorbidities** |  |  |  |
| Medical Comorbidities:<br>Charlson Score (Mean ± SD) | 0.84 ± 1.32 | 1.48 ± 1.72 | 1.26 ± 1.62 |
| Clinical Frailty Score<br>(Intermediate to High Risk) | 171 (5.1%) | 1,160 (18.7%) | 1,331 (13.9%) |
| Heart Failure | 1,023 (30.6%) | 3,775 (60.7%) | 4,798 (50.2%) |
| Coronary Artery Disease | 1,460 (43.7%) | 3,802 (61.1%) | 5,262 (55.0%) |
| Cardiac Arrhythmia | 238 (7.1%) | 1,250 (20.1%) | 1,488 (15.6%) |
| Peripheral Vascular Disease | 25 (0.7%) | 146 (2.3%) | 171 (1.8%) |
| Cerebrovascular Disease | 60 (1.8%) | 227 (3.7%) | 287 (3.0%) |
| Diabetes | 1,137 (34.0%) | 2,568 (41.3%) | 3,705 (38.8%) |
| Hypertension | 2,575 (77.0%) | 5,708 (91.8%) | 8,283 (86.6%) |
| Dyslipidemia | 1,589 (47.5%) | 3,687 (59.3%) | 5,276 (55.2%) |
| Chronic Obstructive Pulmonary Disease | 309 (9.2%) | 1,025 (16.5%) | 1,334 (14.0%) |
| Renal Disease | 59 (1.8%) | 398 (6.4%) | 457 (4.8%) |
| Dialysis | 46 (1.4%) | 192 (3.1%) | 238 (2.5%) |
| Major Neurocognitive Disorder | 36 (1.1%) | 417 (6.7%) | 453 (4.7%) |
| Cancer | 133 (4.0%) | 473 (7.6%) | 606 (6.3%) |
| Liver Disease | 33 (1.0%) | 117 (1.9%) | 150 (1.6%) |
| Interstitial Lung Disease | 11 (0.3%) | 75 (1.2%) | 86 (0.9%) |
| **Prior Cardiac Procedures** |  |  |  |
| Prior Percutaneous Coronary Intervention | 275 (8.2%) | 1,832 (29.5%) | 2,107 (22.0%) |
| **Fiscal Year** |  |  |  |
| 2017 | 812 (24.3%) | 719 (11.6%) | 1,531 (16.0%) |
| 2018 | 847 (25.3%) | 1,015 (16.3%) | 1,862 (19.5%) |
| 2019 | 686 (20.5%) | 1,350 (21.7%) | 2,036 (21.3%) |
| 2020 | 542 (16.2%) | 1,463 (23.5%) | 2,005 (21.0%) |
| 2021 | 455 (13.6%) | 1,671 (26.9%) | 2,126 (22.2%) |

Important temporal trends were noted. The annual number of TAVR procedures increased steadily from 719 to 1,671 from 2017 to 2021, while the annual number of isolated SAVR procedures declined from 812 to 455 in the same time frame.

The proportion of patients in each quintile of the ON-MARG dimensions is displayed in Figure 2. In the residential instability domain, TAVR and SAVR had a higher proportion of patients in quintiles 3 to 5 (more marginalized) and a lower proportion of patients in quintiles 1 and 2 (less marginalized). For ethnic marginalization, TAVR and SAVR both had a higher proportion of patients in quintiles 1 to 3 and a lower proportion of patients in quintiles 4 and 5. For material deprivation, TAVR and SAVR have approximately an equal distribution of patients across all quintiles.

**Figure 2:**
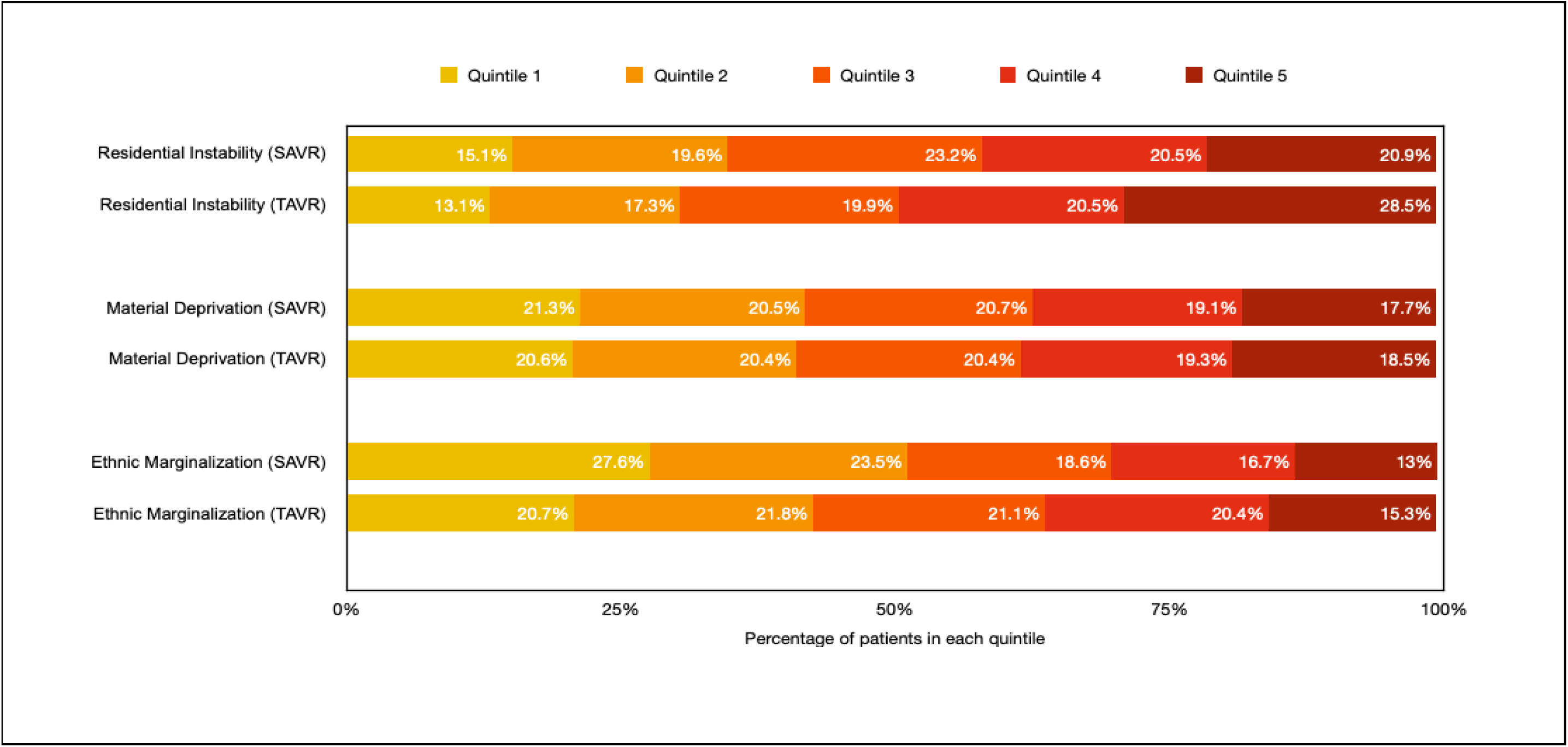
Proportion of patients per quintile by Ontario Marginalization Index dimension for SAVR and TAVR. TAVR, transcatheter aortic valve replacement SAVR, surgical aortic valve replacement”

### Unadjusted Outcomes

Unadjusted outcomes are shown in Table 3. TAVR was associated with higher mortality and higher readmission rates at both 30 days and 1 year post-procedure, compared to SAVR. SAVR had 30-day mortality rates of 1.3% compared to 2.1% for TAVR (p-value = 0.008), while 1-year mortality for SAVR was 3.7% compared to 10.1% for TAVR (p-value < 0.001). 30-day readmission rates for SAVR were 9.8% compared to 11.5% for TAVR (p-value = 0.014); 1-year readmission rates for SAVR were 23.1% compared to 38% for TAVR (p-value < 0.001).

**Table 3:** 30-day and 1-year mortality and hospitalizations after SAVR and TAVR. TAVR, transcatheter aortic valve replacement SAVR, surgical aortic valve replacement

|  | SAVR (N=3,342) | TAVR (N=6,218) | Total (SAVR & TAVR)<br>(N=9,560) | P-value |
| --- | --- | --- | --- | --- |
| Mortality within 30 days after procedure | 44 (1.3%) | 129 (2.1%) | 173 (1.8%) | 0.008 |
| Mortality within 1 year after procedure | 123 (3.7%) | 628 (10.1%) | 751 (7.9%) | <0.001 |
| All-cause hospital readmission from procedure discharge within 30 days | 328 (9.8%) | 712 (11.5%) | 1,040 (10.9%) | 0.014 |
| All-cause hospital readmission from procedure discharge within 1 year | 771 (23.1%) | 2,365 (38.0%) | 3,136 (32.8%) | <0.001 |

### Adjusted Mortality and Readmission Outcomes

Figure 3 depicts adjusted hazard ratios for mortality and readmission at 30 days and 1-year for TAVR-SAVR. The adjusted hazard ratios were 0.58 (95% CI [0.37, 0.92]; p= 0.02) for 30-day mortality and 1.20 (95% CI [0.94, 1.53]; p=0.14) for 1-year mortality, 0.75 (95% CI [0.63, 0.89]; p-value = 0.001) for 30-day readmission and 1.14 (95% CI [1.02, 1.27]; p-value = 0.01) for 1-year readmission. This data shows that, after adjustment for underlying comorbidities, TAVR is associated with lower mortality and readmission at 30 days and higher readmission at 1 year compared to SAVR. There is no significant difference in mortality at 1 year between TAVR and SAVR.

**Figure 3:**
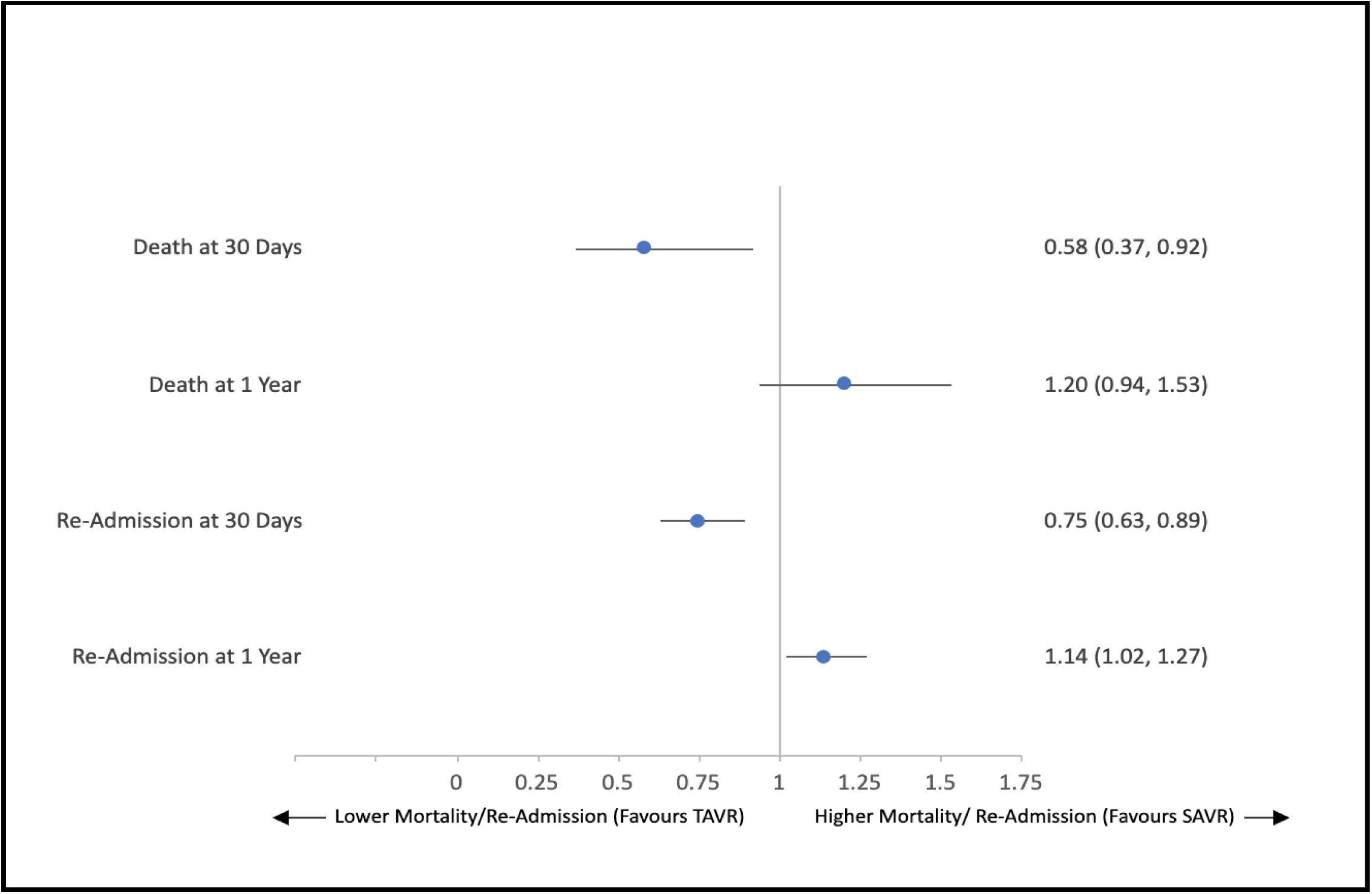
Adjusted hazard ratios for mortality and re-admission at 30 days and 1 year for TAVR vs. SAVR without interactions across the ON-MARG dimensions. The hazard ratio has been adjusted for the following variables: 1) Age 2) Charlson score 3) Frailty score 4) Female sex 5) Rural residence 6) Congestive heart failure 7) Coronary artery disease 8) Atrial arrhythmias 9) Peripheral vascular disease 10) Cerebrovascular disease 11) Diabetes 12) Hypertension 13) Dyslipidemia 14) Chronic obstructive pulmonary disease 15) Major neuroognitive disorder 16) Cancer 17) Renal disease 18) Dialysis 19) Interstitial lung disease 20) Liver disease 21) Prior percutaneous coronary intervention 24) Fiscal year (2017/2018/2019/2020/2021) TAVR, transcatheter aortic valve replacement SAVR, surgical aortic valve replacement

Figure 4 shows the adjusted hazard ratios for 30-day and 1-year mortality and readmissions for TAVR and SAVR, when interactions across the three ON-MARG dimensions are included in the analysis. Across the different quintiles of each ON-MARG domain, there were broadly consistent relationships between TAVR-SAVR and clinical outcomes. For example, TAVR was associated with lower mortality at 30 days for quintiles 3 and 4 for residential instability, quintiles 2 and 4 for material deprivation and quintiles 2 and 5 for ethnic concentration. TAVR was also associated with lower 30-day readmission, compared to SAVR, for quintiles 3 and 5 for residential instability, quintiles 2 and 4 for material deprivation and quintiles 3 and 4 for ethnic concentration. TAVR was associated with a higher 1-year readmission rate for quintile 2 in the residential instability domain and quintiles 1 and 3 in the material deprivation domain. There were no associations between the three ON-MARG dimensions and mortality at 1-year post-procedure. Given these findings, the main effect of TAVR-SAVR on mortality and re-admission was consistent regardless of ethnicity, material deprivation and residential instability.

**Figure 4:**
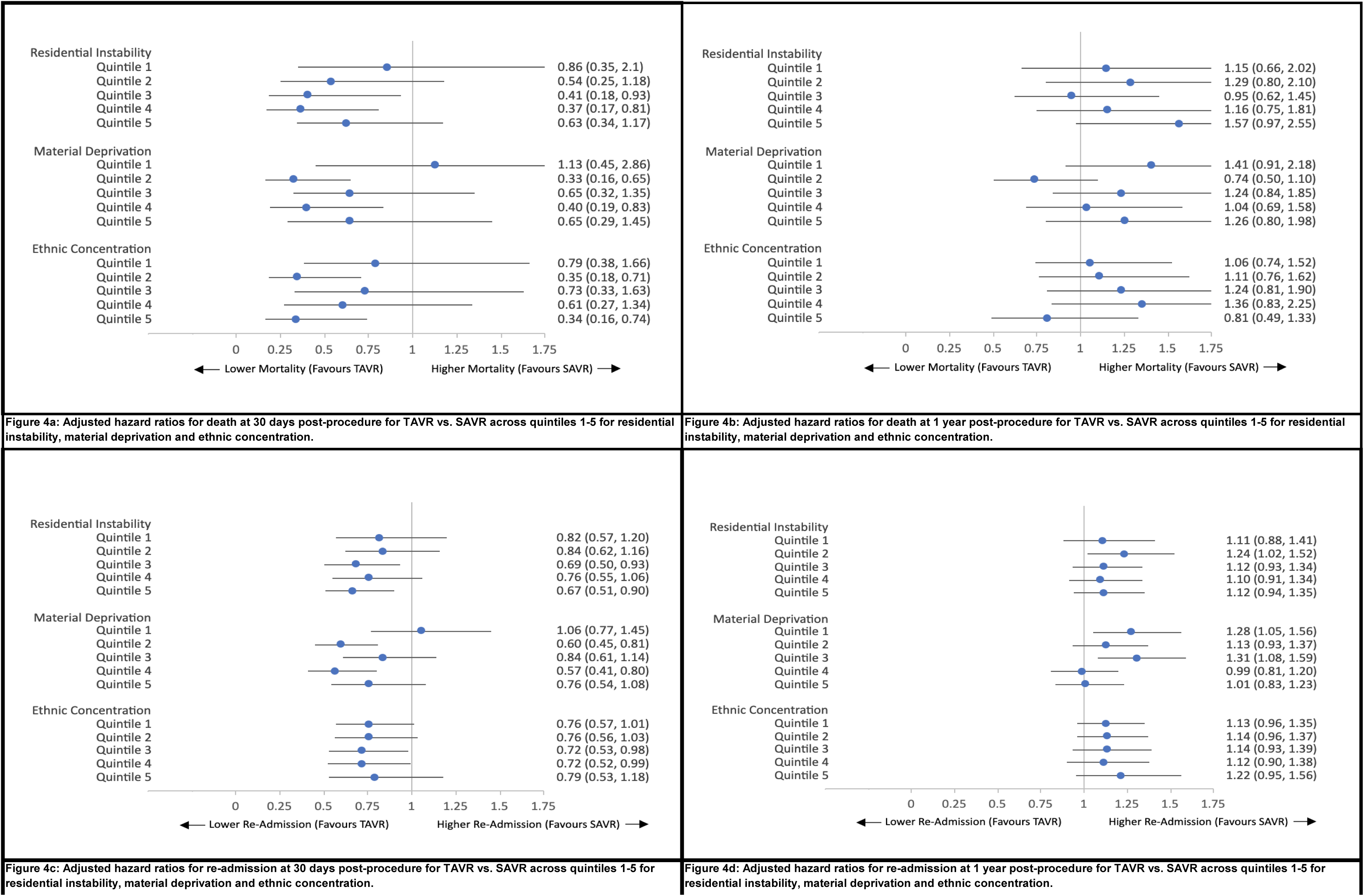
The hazard ratio has been adjusted for the following variables: 1) Age 2) Charlson score 3) Frailty score 4) Female sex 5) Rural residence 6) Congestive heart failure 7) Coronary artery disease 8) Atrial arrhythmias 9) Peripheral vascular disease 10) Cerebrovascular disease 11) Diabetes 12) Hypertension 13) Dyslipidemia 14) Chronic obstructive pulmonary disease 15) Major neuroognitive disorder 16) Cancer 17) Renal disease 18) Dialysis 19) Interstitial lung disease 20) Liver disease 21) Prior percutaneous coronary intervention 24) Fiscal year (2017/2018/2019/2020/2021) TAVR, transcatheter aortic valve replacement SAVR, surgical aortic valve replacement

## DISCUSSION

The objective of our study was to investigate the relationship between social deprivation and outcomes after TAVR and SAVR. The key findings from this study are as follows:

**1.** TAVR is associated with a lower adjusted mortality rate and readmission rate at 30 days and a higher readmission rate at 1 year compared to SAVR.
**2.** The association between TAVR and SAVR and these outcomes is irrespective of markers of social deprivation, including ethnic concentration, residential instability and material deprivation.

Prior to our study, there was limited data evaluating the impact of social deprivation on outcomes after both novel and established cardiac procedures. A retrospective analysis in the United Kingdom evaluated initial and long-term outcomes in all patients who underwent PCI and/or CABG between April 2007 and March 2012 and determined the association between these outcomes and levels of social deprivation using an established index of multiple deprivation (IMD) scores^6^. After appropriate baseline risk factor adjustment, there were significant associations between social deprivation and length of hospitalization, as well as mortality for patients undergoing PCI alone. There was no association between social deprivation and outcomes for patients undergoing CABG alone or PCI with CABG^6^. In the United States, a retrospective analysis of 17,973 TAVR patients and 95,078 SAVR patients was conducted to determine the relationship between racial status and procedural outcomes^9^. After appropriate adjustment, Black patients had higher 30-day re-admission rates after SAVR compared to White patients, but there were no differences in mortality. There were no race-related differences in mortality or re-admission after TAVR^9^. Apart from these two retrospective studies, there are no other studies evaluating the association between social deprivation and comparative outcomes after newer and established cardiac procedures. Additionally, there is limited data comparing the outcomes of these procedures and their association with other markers of social deprivation, including material deprivation and residential instability, which both may represent a complex interplay with ethnicity.

Although our study was not designed to understand the mechanism between the various markers of social deprivation and outcomes after TAVR and SAVR, we can hypothesize several plausible explanations for our findings. Residential instability is known to be associated with increased emergency department service utilization, subsequent repeat hospitalizations, as well as mortality and multimorbidity^12^. A recent study showed that residential instability was associated with increased rates of 1-year mortality after TAVR^8^. Although there are no specific studies evaluating the impact of residential instability on outcomes after SAVR, it can be hypothesized that housing instability impacts follow-up, subsequent emergency department visits, re-hospitalization and mortality in a similar way post-SAVR and post-TAVR.

The interaction mechanism between ethnic concentration and post-procedural outcomes is more challenging to describe. A prior study showed that outcomes after TAVR do not seem to be associated with ethnic concentration^8^. Additionally, based on the ON-MARG classification criteria, ethnic concentration scores account for both recent immigrants and visible minorities. In this context, the healthy immigrant phenomenon may bias the analysis, as new immigrants tend to be healthier than average Canadians, leading to lower rates of post-procedural adverse outcomes and re-hospitalization^13,14^. Therefore, as ON-MARG’s definition of ethnic concentration describes a diverse population subtype, the results achieved in our analysis are not a simple representation of race-related impacts on procedural outcomes. This in turn may obscure any differential impact on TAVR versus SAVR.

The impact of material deprivation on procedural outcomes is also not intuitive. It has been described that, in its initial growth phase, TAVR was more likely to be established in hospitals serving wealthier patients, leading to inequities in access to TAVR in poorer communities^15^. Similarly, it can be hypothesized that other cardiac programs, such as SAVR, are also more likely to be established first in these wealthier settings. In this context, patients with higher levels of material deprivation may have less access to either of these procedures.

Additionally, we found that, after adjustment for appropriate clinical and demographic variables, TAVR was associated with a lower mortality rate at 30 days compared to SAVR, regardless of any social deprivation measures. This finding is in keeping with prior randomized controlled trials, where TAVR was associated with a reduction in 30-day mortality compared to SAVR in both high-risk and low-risk surgical candidates^2,4^. A recent observational study of 33,789 isolated TAVR and SAVR procedures in Germany found that after risk factor adjustment, in patients that were older than 80 years with NYHA class 3 or 4 heart failure or in patients with renal failure, TAVR was associated with lower in-hospital mortality compared to SAVR^16^. Similar to these studies, our patient population consisted of an intermediate-higher risk patient group with high rates of concurrent heart failure, coronary artery disease, cardiac arrhythmias and high clinical frailty scores, which likely explains the association between lower 30-day mortality with TAVR compared to SAVR in our study.

## LIMITATIONS

There are several limitations to this study. This study is a retrospective observational study, which may be affected by unknown and unmeasured confounders that were not incorporated into our model and may impact our outcome analysis. We used administrative databases in our study, which are limited by the accuracy of administrative codes to track procedural outcomes. The ON-MARG is a proxy for social marginalization and may not account for the heterogeneity that exists on an individual level, as quintiles are distributed over large geographic areas. It is also important to acknowledge that only patients who were referred to TAVR and SAVR were included in the analysis, and patients who did not have access to these referrals or cardiac care were excluded from the analysis altogether. Finally, this is a study that was conducted in Ontario, Canada and may not be generalizable to other regions, including jurisdictions without universal healthcare systems. Indeed, it would be useful to conduct future studies in other jurisdictions, such as for example in the United States, with alternative approaches to health care delivery, to determine whether there is a more apparent impact of social deprivation on post-procedural outcomes, contrasting between the type of procedure (TAVR versus SAVR).

## CONCLUSION

In conclusion, our study found that individual markers of social deprivation, namely ethnic concentration, material deprivation and residential instability, did not impact short-term or long-term hospitalization and mortality outcomes after TAVR and SAVR. TAVR is associated with lower 30-day mortality and readmission and higher 1-year re-admission rates post-procedure, compared to SAVR, and these associations exist irrespectively of the three markers of social deprivation. Further studies are warranted to test these associations across different novel and established cardiac procedures and different jurisdictions, as interactions between markers of social deprivation and procedural outcomes are essential in influencing economic and health policy decisions.

## ACKNOWLEDGEMENTS

This study was supported by ICES, which is funded by an annual grant from the Ontario Ministry of Health (MOH) and the Ministry of Long-Term Care (MLTC). This document used data adapted from the Statistics Canada Postal Code^OM^ Conversion File, which is based on data licensed from Canada Post Corporation, and data adapted from the Ontario Ministry of Health Postal Code Conversion File, which contains data copied under license from ©Canada Post Corporation and Statistics Canada. Parts of this material are based on data and information compiled and provided by the Ontario Ministry of Health. The analyses, conclusions, opinions and statements expressed herein are solely those of the authors and do not reflect those of the funding or data sources; no endorsement is intended or should be inferred. Parts of this material are based on data and information compiled and provided by: the Canadian Institute of Health Information (CIHI) and CorHealth Ontario. The analyses, conclusions, opinions and statements expressed herein are solely those of the authors and do not reflect those of the funding or data sources; no endorsement is intended or should be inferred. Parts of this material are based on data and/or information compiled and provided by CIHI. However, the analyses, conclusions, opinions and statements expressed in the material are those of the authors, and not necessarily those of CIHI. The authors acknowledge that the clinical registry data used in this publication is from participating hospitals through CorHealth Ontario, which serves as an advisory body to the MOH, is funded by the MOH, and is dedicated to improving the quality, efficiency, access and equity in the delivery of the continuum of adult cardiac, vascular and stroke services in Ontario, Canada. We thank IQVIA Solutions Canada Inc. for use of their Drug Information File.

We thank the Toronto Community Health Profiles Partnership for providing access to the Ontario Marginalization Index. The corresponding author affirms that he has listed everyone who contributed significantly to the work. The authors had access to all the study data, take responsibility for the accuracy of the analysis, and had authority over manuscript preparation and the decision to submit the manuscript for publication. The corresponding author confirms that all authors read and approve the manuscript.

## CITATIONS

1. Makkar, R. R., Fontana, G. P., Jilaihawi, H., Kapadia, S., Pichard, A. D., Douglas, P. S., … & Leon, M. B. (2012). Transcatheter aortic-valve replacement for inoperable severe aortic stenosis. New England Journal of Medicine, 366(18), 1696–1704.

2. Smith, C. R., Leon, M. B., Mack, M. J., Miller, D. C., Moses, J. W., Svensson, L. G., … & Pocock, S. J. (2011). Transcatheter versus surgical aortic-valve replacement in high-risk patients. New England Journal of Medicine, 364(23), 2187–2198.

3. Leon, M. B., Smith, C. R., Mack, M. J., Makkar, R. R., Svensson, L. G., Kodali, S. K., … & Webb, J. G. (2016). Transcatheter or surgical aortic-valve replacement in intermediate-risk patients. New England Journal of Medicine, 374(17), 1609–1620.

4. Mack, M. J., Leon, M. B., Thourani, V. H., Makkar, R., Kodali, S. K., Russo, M., … & Smith, C. R. (2019). Transcatheter aortic-valve replacement with a balloon-expandable valve in low-risk patients. New England Journal of Medicine, 380(18), 1695–1705.

5. Matheson FI, Moloney G, van Ingen T; Ontario Agency for Health Protection and Promotion (Public Health Ontario). 2016 Ontario marginalization index: user guide. 1st revision. Toronto, ON: St. Michael’s Hospital (Unity Health Toronto); 2022. Joint publication with Public Health Ontario.

6. Matata, B. M., Shaw, M., Grayson, A. D., McShane, J., Lucy, J., Fisher, M., & Jackson, M. (2016). The impact of social deprivation on coronary revascularisation treatment outcomes within the National Health Service in England and Wales. European Journal of Preventive Cardiology, 23(3), 316–327.

7. Barnard, J., Grant, S. W., Hickey, G. L., & Bridgewater, B. (2015). Is social deprivation an independent predictor of outcomes following cardiac surgery? An analysis of 240 221 patients from a national registry. BMJ open, 5(6), e008287.

8. Patel, R. V., Ravindran, M., Qiu, F., Manoragavan, R., Sud, M., Tam, D. Y., … & Wijeysundera, H. C. (2023). Social Deprivation and Post-TAVR Outcomes in Ontario, Canada: A Population-Based Study. Journal of the American Heart Association, 12(1), e028144.

9. McNeely, C., Zajarias, A., Fohtung, R., Kakouros, N., Walker, J., Robbs, R., … & Vassileva, C. M. (2018). Racial comparisons of the outcomes of transcatheter and surgical aortic valve implantation using the Medicare database. The American Journal of Cardiology, 122(3), 440–445.

10. Taylor, N. E., O’Brien, S., Edwards, F. H., Peterson, E. D., & Bridges, C. R. (2005). Relationship between race and mortality and morbidity after valve replacement surgery. Circulation, 111(10), 1305–1312.

11. Wijeysundera, H. C., Qiu, F., Koh, M., Prasad, T. J., Cantor, W. J., Cheema, A., … & Ko, D. T. (2017). Comparison of outcomes of balloon-expandable versus self-expandable transcatheter heart valves for severe aortic stenosis. The American Journal of Cardiology, 119(7), 1094–1099.

12. Zygmunt, A., Tanuseputro, P., James, P., Lima, I., Tuna, M., & Kendall, C. E. (2020). Neighbourhood-level marginalization and avoidable mortality in Ontario, Canada: a population-based study. Canadian Journal of Public Health, 111, 169–181.

13. Ali, J. S., McDermott, S., & Gravel, R. G. (2004). Recent research on immigrant health from Statistics Canada’s population surveys. Canadian Journal of Public Health, 95, I9–I13.

14. Omariba, D. W. R. (2015). Immigration, ethnicity, and avoidable mortality in Canada, 1991–2006. Ethnicity & health, 20(4), 409–436.

15. Nathan, A. S., Yang, L., Yang, N., Khatana, S. A. M., Dayoub, E. J., Eberly, L. A., … & Fanaroff, A. C. (2021). Socioeconomic and geographic characteristics of hospitals establishing transcatheter aortic valve replacement programs, 2012–2018. Circulation: Cardiovascular Quality and Outcomes, 14(11), e008260.

16. Stachon, P., Kaier, K., Zirlik, A., Bothe, W., Heidt, T., Zehender, M., … & von Zur Mühlen, C. (2019). Risk-adjusted comparison of in-hospital outcomes of transcatheter and surgical aortic valve replacement. Journal of the American Heart Association, 8(7), e011504.

